# Triangulating evidence to evaluate selection bias in lifecourse Mendelian randomization studies: a practical framework illustrated by early-life adiposity and breast cancer

**DOI:** 10.1101/2025.09.10.25335479

**Authors:** Grace M. Power, Eleanor Sanderson, Apostolos Gkatzionis, Tom G. Richardson, Kate Tilling, George Davey Smith, Gibran Hemani

## Abstract

**Background:** Higher adiposity in early-life has consistently been associated with a reduced risk of breast cancer in later life, with Mendelian randomization (MR) studies supporting a causal effect. However, concerns have been raised that selection bias, particularly collider stratification due to selective participation or survival, may induce spurious protective MR estimates.

**Methods:** We triangulated across empirical analyses and simulations to evaluate whether selection-induced bias could plausibly explain the inverse effect estimate of early-life adiposity on breast cancer risk. First, we analysed proxy-genotype Mendelian randomization (MR) analyses of breast cancer in relatives, in which participant genotype is used as a proxy for relatives’ genotype, and conducted family-based simulations to assess whether attenuation in relative-based estimates could arise without selection bias. Second, we performed multivariable MR analyses of parental survival to evaluate survival-related selection mechanisms. Third, we conducted extensive simulations to quantify the magnitude of bias introduced by selection under a range of plausible and extreme scenarios, including interaction-driven selection.

**Results:** The weaker proxy-genotype MR estimates of breast cancer in relatives, compared with MR estimates for an individual’s own breast cancer, were reproduced in family-based simulations without selection bias, indicating that this pattern does not provide evidence for selection bias. Multivariable MR analyses of parental survival indicated that survival differences are primarily driven by mid-to-late adulthood, not early-life, adiposity, providing little support for survival-related selection acting through early-life adiposity. In simulation analyses, additive selection produced minimal bias, while interaction-driven selection generated increasing distortion; however, even under extreme scenarios, the magnitude of bias was insufficient to replicate the observed protective effect estimate. In simulations where selection depended on mid-to-late adulthood rather than early-life adiposity, bias was expressed primarily in mid-to-late adulthood MR estimates, with little distortion of early-life MR estimates. Across all simulated scenarios, the combined pattern of empirical univariable and multivariable MR findings was not reproduced by selection alone.

**Conclusions:** Although selection bias can influence MR estimates, our findings suggest that plausible selection mechanisms are unlikely to substantively explain the observed inverse effect estimate of early-life adiposity on breast cancer risk. These results support a causal interpretation of the strong protective effect estimate of early-life adiposity on breast cancer risk and highlight the value of triangulating evidence across complementary approaches when evaluating bias in lifecourse MR.

**Plain Language Summary:** Previous studies have found that having a larger body size in early life is linked to a lower risk of developing breast cancer later in life. Mendelian randomization studies, which use genetic variation to investigate possible causal effects, have supported this finding. However, it has been suggested that the result could be explained by selection bias arising from who survives or participates in the studies analysed. We used several complementary approaches to evaluate this possibility. Together, the findings provide little evidence that the selection mechanisms examined explain the protective effect estimate of early-life adiposity on breast cancer. This work also provides a practical framework for investigating selection bias in lifecourse Mendelian randomization studies.

**What is new?:**

1. Key findings
  - Proxy-genotype Mendelian randomization analyses indicate that attenuation in relative-based estimates is explained by Mendelian inheritance and measurement dilution rather than selection bias.
  - Parental survival analyses provide little support for survival-related selection acting independently through early-life adiposity as an explanation for its protective effect on breast cancer.
  - Simulation studies show that even extreme selection mechanisms do not reproduce the magnitude or overall pattern of empirical lifecourse Mendelian randomization effect estimates for early-life adiposity and breast cancer.
2. What this adds to what is known?
  - Triangulating complementary empirical analyses with targeted simulations provides a practical framework for evaluating whether proposed selection mechanisms are sufficient to explain findings from lifecourse Mendelian randomization studies.
3. What is the implication, what should change now?
  - Concerns about selection bias should be evaluated using multiple complementary lines of evidence rather than assumed to invalidate findings from lifecourse Mendelian randomization studies.

## 1. Background

Higher levels of adiposity in early life have consistently been observed to be inversely associated with breast cancer risk later in life across multiple lines of evidence [1–12]. Mendelian randomization (MR) studies, which use germline genetic variants as instrumental variables, strengthen the case for a causal interpretation by mitigating confounding and reverse causation [13]. In univariable MR analyses, both higher earlier life and higher mid-to-late adulthood adiposity have been estimated to exert protective causal effect estimates on breast cancer risk [1–6]. In lifecourse MR applications, causal effect estimates of the same trait measured at different life stages can be separated using a multivariable MR framework [14, 15]. Within this framework, the apparent protective effect of mid-to-late adulthood adiposity attenuates, whereas the direct effect of early-life adiposity persists after adjustment [1–6].

These findings have prompted debate about whether selection processes could induce spurious protective estimates under a null causal model [16, 17]. These critiques of the early-life adiposity-breast cancer findings are framed primarily in terms of collider stratification bias, whereby conditioning on selection influenced by both early-life adiposity (or its causes) and breast cancer risk (or its causes) can induce a non-causal estimate, even if the true causal effect is null. Accordingly, here we use the term selection bias to refer specifically to this collider-driven mechanism, while recognising that selection bias can arise through other forms of selection or stratification [18, 19]. The magnitude of such collider-induced bias depends on the underlying causal structure and interactions influencing selection, and may be underestimated if these are ignored [20, 21]. Bias arising from selection on an effect modifier (i.e., when study participation depends on a variable that changes the size of the exposure effect), which can occur even in the absence of collider structures [22], is not considered here. This is because it requires the exposure to have a causal effect and therefore cannot operate under the sharp null hypothesis (i.e., where the causal effect is zero for every individual) and is not relevant to the present evaluation [23].

Since early methodological discussions of selection bias in epidemiology, it has been emphasised that the possibility of selection bias should not be a barrier to future research but should instead motivate careful evaluation of whether plausible selection mechanisms are sufficient to explain observed findings [18]. Rather than assuming that selection bias necessarily invalidates lifecourse MR results, this commentary uses triangulation across several complementary empirical and simulation-based approaches to evaluate whether selection-induced collider stratification is sufficient to explain the observed protective effect estimate. Data sources for empirical analyses are described in Note S1; Table S1.

## 2. Proxy-genotype Mendelian randomization (MR) analyses

Work using proxy-genotype MR has been used to argue that the protective effect estimate of early-life adiposity on breast cancer may be explained by selection bias [16, 17]. In these analyses, the participant’s genotype is used as a proxy for the genotype of relatives, allowing breast cancer occurring in mothers or siblings, including diagnoses before participant recruitment, to be analysed. Because the estimated effect is weaker for relatives than for an individual’s own breast cancer, this attenuation has been interpreted as evidence that the observed protective effect estimate may be partly explained by selection bias. The rationale is that breast cancer diagnoses in relatives can occur before participant recruitment and are therefore less susceptible to survival-related selection at recruitment. However, attenuation is expected even in the absence of selection bias because the participant’s genotype is only an imperfect proxy for the genotype of relatives owing to Mendelian inheritance, resulting in exposure measurement error dilution [11].

We conducted a family-based simulation in which early-life adiposity had the same causal effect estimate on breast cancer in mothers, siblings and probands. Within the simulated data, we re-performed the summary-based MR analysis to estimate the effect of early-life adiposity on own breast cancer, as well as the proxy-MR estimates of own early-life adiposity on sibling’s breast cancer and mother’s breast cancer. Self- and proxy-MR estimates were then obtained from the simulation data. No selection bias was introduced into the simulation framework. We show that the observed pattern of estimates for self, sibling, and mother designs is reproduced within the basic proxy-MR framework and does not require selection bias to explain it (Figure S1).

## 3. Evaluating survival-related selection via parental survival analyses

A further line of critique has suggested that the apparent protective effect estimate of early-life adiposity on breast cancer could arise through survival-related selection, whereby early-life adiposity influences mortality prior to recruitment [16, 17]. For the proposed survival-selection mechanism to explain the reported protective effect estimate on breast cancer, early-life adiposity would need to have an effect on survival that is independent of mid-to-late adulthood adiposity. However, early-life and mid-to-late adulthood adiposity are highly genetically correlated, and mid-to-late adulthood adiposity has a well-established adverse effect on survival. Consequently, any apparent effect estimate of early-life adiposity on survival could simply reflect the effect of mid-to-late adulthood adiposity rather than an independent effect of early-life adiposity. To distinguish these effects, we conducted multivariable MR analyses using the UK Biobank data described in Note S1, applying the same early-life and mid-to-late adulthood adiposity genetic instruments used by Richardson et al. [1]. We used parental survival in the absence of uncensored individual survival measures as the outcome because parental lifespan strongly predicts participants’ own lifespan at the population level, yet is unlikely to be substantially causally influenced by the participant’s own early-life or mid-to-late adulthood adiposity (Note S2). If the apparent protective effect of early-life adiposity on breast cancer arose primarily from survival-related selection acting through the genetic instruments, early-life adiposity would also be expected to retain an independent effect on parental survival after accounting for mid-to-late adulthood adiposity. We therefore estimated the direct effect of early-life adiposity on parental survival after accounting for mid-to-late adulthood adiposity.

In MR analyses of parental survival, with offspring genotype used as a proxy for parental genotype, we found that higher mid-to-late adulthood adiposity exerted a strong adverse effect on survival in both univariable (OR = 0.66, 95% CI: 0.62 to 0.71; P = 1.66 x 10-37) and multivariable MR analyses (OR = 0.67, 95% CI: 0.62 to 0.73; P = 2.33 x 10-21), whereas the effect estimate for early-life adiposity observed in univariable analyses (OR = 0.80, 95% CI: 0.75 to 0.86; P = 4.49 x 10-12) fully attenuated after adjustment for mid-to-late adulthood adiposity (OR = 1.00, 95% CI: 0.91 to 1.10; P = 0.955; Table S2). These findings indicate that the apparent effect of early-life adiposity on survival is largely explained by its correlation with mid-to-late adulthood adiposity, rather than an independent effect of early-life adiposity. This suggests that survival-related selection acting through early-life adiposity is unlikely to explain the protective effect on breast cancer risk. Full results are provided in Table S2. This interpretation is further supported by observational studies suggesting that the protective effect of early-life adiposity on breast cancer is not simply explained by later mid-to-late adulthood adiposity [7, 12].

## 4. Evaluating selection-induced collider stratification

Selection mechanisms can induce and distort effect estimates, and this is a particular concern in settings such as the UK Biobank, where participation is selective (5.5% response rate) and recruitment occurs in mid-to-late adulthood [24]. The relevant question, however, is whether plausible selection mechanisms could generate effect estimates of the magnitude and pattern observed in lifecourse MR. We therefore conducted simulations to evaluate this possibility.

Because of the scale and two-sample design of the empirical lifecourse MR analyses, reproducing selection mechanisms that perfectly match observed parameters is not easily tractable. Instead, we used simulations to evaluate whether collider-based selection processes, operating with or without interaction, could reproduce the magnitude and pattern of lifecourse MR estimates reported for early-life adiposity and breast cancer (Table S3). Data were generated under a sharp null model, assuming no true causal effect of either early-life or mid-to-late adulthood adiposity on breast cancer risk for any individual. The simulations are intended to be illustrative rather than definitive, with full methodological details and results provided in the Supplementary Material.

To inform the specification of the simulations, we first examined the influence of confounding in conventional observational analyses (Note S3). In minimally adjusted logistic regression analyses of UK Biobank data, higher early-life adiposity remained inversely associated with breast cancer risk after adjustment for mid-to-late adulthood adiposity and age. The magnitude of this association was smaller than that observed in the MR analyses (Note S4). This is consistent with attenuation of observational estimates arising from measurement error in retrospectively reported early-life adiposity, to which instrumental variable analyses are less susceptible to under standard assumptions. However, residual confounding could still contribute to the observational association. Accordingly, we included an unmeasured confounder in the simulation framework to reflect this empirically informed structure.

### 4.1 Magnitude of bias that could arise under simulated selection mechanisms

To assess the strength of selection required to reproduce the magnitude of the protective lifecourse MR estimate under a null causal model, we conducted simulations in which early-life and mid-to-late adulthood adiposity were modelled as latent continuous traits instrumented by correlated genetic risk scores (Note S5). Adult adiposity was specified to depend partly on early-life adiposity, reflecting the tendency for adiposity to persist across the lifecourse. Breast cancer status was generated independently of adiposity, such that the true causal effects of early-life and mid-to-late adulthood adiposity were set to zero; any estimated effects therefore reflect bias introduced by selection.

Selection into the analytic sample was modelled as a function of early-life adiposity and breast cancer status, with and without interaction (Figure 1A). We examined 27 scenarios spanning no selection, additive selection, and in e action-driven selection, with parame ers varied from the null to magnitudes exceeding those typically observed empirically, in order to assess upper-bound collider bias (Tables S5 and S6). Lifecourse MR analyses were performed in samples corresponding to the number of female UK Biobank participants with complete data (n = 246,511), with 500 simulation replicates per setting.

**Figure 1.**
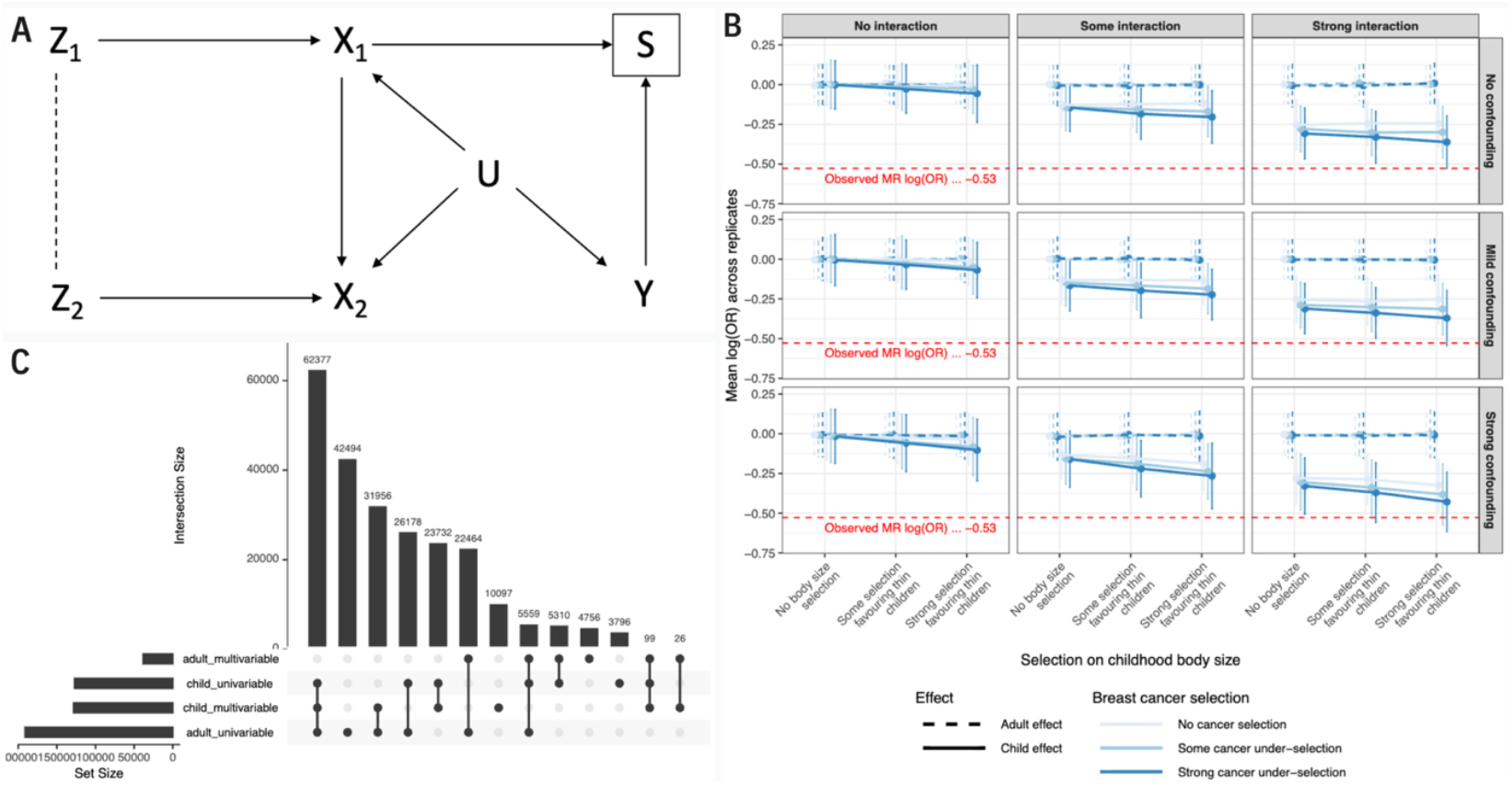
Schematic of the simulated selection mechanism and resulting bias under varying selection and unmeasured confounding scenarios. A. Early-life (***Z1***) and mid-to-late adulthood (***Z2***) genetic risk scores (GRS) influence latent early-life (***X1***) and mid-to-late adulthood (***X2***) adiposity, respectively, with a genetic correlation (dotted line) between **Z1**and**Z2**. Early-life adiposity (***X1***) and breast cancer status (*Y*) influence selection into the analytic sample (*S*), representing either differential survival prior to recruitment or differential participation among individuals alive at recruitment. An unmeasured confounder (*U*) influences early-life adiposity, mid-to-late adulthood adiposity, and breast cancer risk, allowing assessment of whether conditioning on S induces or amplifies collider bias through non-causal pathways. The strength of confounding was varied across two levels: mild, where *U* was weakly associated with these variables, and strong, where *U* had stronger associations with them. B. Mean log(OR) MR estimates for early-life (solid) and mid-to-late adulthood (dashed) adiposity on breast cancer across 500 replicates are shown under increasing strengths of unmeasured confounding (U) affecting both adiposity and breast cancer risk. Horizontal panels represent different confounding strengths (no, mild, strong), and vertical panels represent increasing levels of interaction-driven selection. Error bars indicate the 2.5^th^-97.5^th^ percentile range. The red dashed line marks the observed early-life MR estimate (logOR =-0.53). C. UpSet plot showing how often simulation scenarios produce results that overlap with empirical results across four MR estimates.

Empirical estimates of disease-related selection into UK Biobank informed the lower range of interaction parameters considered (Note S6; Figure S2; Table S7). For breast cancer in women, cases are slightly over-represented in UK Biobank relative to the general population, with modest age-related interaction. Parameter values in the simulations were however extended beyond these empirical magnitudes, including configurations implying substantial case under-representation, in order to evaluate whether extreme selection structures could generate MR effect estimates comparable to those observed. The simulations thus represent a stress test of the selection hypothesis rather than an attempt to reproduce the precise selection mechanism operating in UK Biobank.

Consistent with the multivariable MR analyses of parental survival, which indicated that survival-related differences are primarily associated with mid-to-late adulthood rather than early-life adiposity, we conducted a sensitivity analysis in which selection was specified to operate through mid-to-late adulthood adiposity and breast cancer status, with early-life selection fixed at zero.

In the primary simulations where selection depended on early-life adiposity and breast cancer status (Figure 1A), additive selection produced little distortion of early-life MR estimates (Table S8). Interaction-dependent selection induced progressively more negative early-life estimates as interaction magnitude increased (Figure 1B), and the presence of confounding amplified this distortion. However, even under the most extreme combinations of interaction and confounding evaluated, simulated early-life estimates remained less extreme than the empirical lifecourse MR estimate (logOR =-0.53).

When selection was instead specified to operate through mid-to-late adulthood adiposity, bias was expressed primarily in mid-to-late adulthood MR estimates, while early-life estimates remained centred near the null across scenarios (Figure S4). This contrast indicates that bias tracks the exposure driving selection. iven that survival-related differences appear to be driven predominantly by mid-to-late adulthood adiposity, a structure in which selection operates primarily through mid-to-late adulthood would be expected to distort mid-to-late adulthood estimates rather than generate a spurious protective early-life effect estimate of the magnitude observed.

Results from simulations without confounding are presented as a baseline in Figure 1B, with coverage of the empirical MR estimate under selection alone shown in Figure S3.

### 4.2 Recapitulating univariable and multivariable MR patterns under simulated selection mechanisms

A further way to evaluate the selection hypothesis is to assess whether simulated selection mechanisms can reproduce the overall pattern of univariable and multivariable MR estimates observed empirically. Specifically, we examined whether selection bias could generate strong protective effect estimates of both early-life and mid-to-late adulthood adiposity on breast cancer in univariable MR, while attenuating only the mid-to-late adulthood adiposity estimate in multivariable MR. To examine this, we extended the previous simulation study to perform multivariable MR. ach simulation therefore produced two univariable MR estimates and two multivariable MR estimates. In addition to simulating no true effect of early-life or mid-to-late adulthood adiposity on cancer, we also included scenarios involving a moderate influence of mid-to-late adulthood adiposity on cancer. ach simulated estimate was compared with its corresponding empirical MR estimate from Richardson et al. (Table S3). For each comparison, we calculated the probability that the simulated and empirical estimates overlapped after accounting for the uncertainty of both estimates (Note S7). A simulation was considered a ‘match’ when this overlap probability exceeded 0.1. Our primary interest was whether all four MR estimates matched simultaneously under a single simulated selection mechanism.

The empirical MR estimates from Richardson et al. served only as reference targets rather than values to be formally calibrated. We again note that a formal sensitivity analysis would seek to parameterise the simulations to match the observed data-generating process [25], which was not our objective here. Across 781,250 simulation scenarios, none reproduced all four empirical MR estimates according to the predefined matching criterion (defined as satisfying the matching criteria above; Figure 1C).

## 5. Discussion

Selection bias is a recognised concern in conventional epidemiology and MR, particularly in large cohort studies with selective participation and recruitment in mid-to-late adulthood. Such concerns warrant careful evaluation of whether plausible selection mechanisms could meaningfully distort effect estimates, rather than presuming that observed effects are artefactual. Triangulating across complementary lines of evidence (Table 1), we examined whether selection-induced collider stratification could plausibly explain the inverse effect estimates of early-life adiposity on breast cancer risk reported in lifecourse MR and conventional epidemiology studies.

**Table 1.** Triangulated evaluation of selection bias as an explanation for the protective effect estimates of early-life adiposity on breast cancer risk.

| Approach | Selection mechanism evaluated | Observed pattern for breast cancer | Implication for selection hypothesis |
| --- | --- | --- | --- |
| **Prospective early-life cohort with registry follow-up**<br>(published observational evidence) | Mid-to-late life recruitment selection in adult cohorts | Higher early-life adiposity associated with lower breast cancer incidence when BMI measured prospectively in early-life and cancer identified via registries | Protective association is observed outside adult volunteer cohorts and does not rely on retrospective exposure reporting, reducing the likelihood that adult-cohort participation alone explains the finding |
| **Proxy-genotype MR of breast cancer in relatives and family-based simulation** | Whether attenuation in relative-outcome MR reflects selection bias | MR effect estimates for breast cancer in mothers or siblings are weaker than for own breast cancer; the same attenuation pattern is reproduced in simulations without selection | Attenuation in proxy MR does not uniquely indicate selection bias and therefore does not provide direct evidence that selection explains the early-life MR estimate for breast cancer |
| **Parental survival MR** | Survival-related selection acting through genetic instruments | Mid-to-late adulthood adiposity shows a persistent adverse effect estimate on parental survival after adjustment; the early-life effect estimate attenuates in multivariable models | Survival differences appear primarily related to mid-to-late adulthood adiposity, limiting support for a early-life-driven survival-selection explanation of the breast cancer finding |
| **Simulations - selection operating through early-life adiposity ($X_1$)** | Inclusion depends on early-life adiposity and breast cancer status, with and without interaction | Additive selection produces minimal distortion of early-life MR effect estimates for breast cancer; interaction-dependent selection induces increasing downward bias | Early-life-driven selection requires interaction to generate appreciable distortion of the early-life MR estimate for breast cancer |
| **Simulations - selection operating through mid-to-late adulthood adiposity ($X_2$)** | Inclusion depends on mid-to-late adulthood adiposity and breast cancer status, with and without interaction | Bias is expressed primarily in mid-to-late adulthood MR effect estimates for breast cancer; early-life estimates remain close to null | When selection depends on mid-to-late adulthood adiposity, bias affects mid-to-late adulthood MR estimates rather than the early-life effect estimates for breast cancer |
| **Extended simulations of univariable and multivariable MR pattern - selection operating through early-life adiposity ($X_1$)** | Whether broader interaction structures can reproduce the empirical lifecourse MR pattern for breast cancer | The joint empirical pattern of early-life and mid-to-late adulthood MR effect estimates for breast cancer is not reproduced under simulated selection alone | The lifecourse MR pattern for breast cancer is not easily recapitulated by selection mechanisms under a null causal model |

An important line of evidence comes from a prospective early-life cohort in which BMI was measured directly in school health examinations and participants were followed through linkage to national cancer registries, which demonstrated similar protective associations [7, 10]. Because exposure was recorded in early-life and outcomes were ascertained through population-based registry linkage rather than voluntary adult recruitment, these findings are less susceptible to participation mechanisms specific to adult cohort studies.

Genetically informed analyses provide further context. Attenuation observed in proxy-genotype MR analyses of breast cancer in relatives is consistent with Mendelian inheritance and measurement dilution and can be reproduced in family-based simulations without introducing selection bias. MR analyses of parental survival indicate that survival-related differences associated with adiposity are primarily attributable to mid-to-late adulthood rather than early-life adiposity, limiting support for a early-life-driven survival selection explanation of the breast cancer finding.

Simulation analyses conducted under a sharp null model assessed the magnitude of bias that could arise under explicit selection structures. Additive selection on early-life adiposity and breast cancer status produced little distortion of early-life MR estimates. Interaction parameters were varied beyond typical empirical magnitudes to approximate upper-bound selection effects. Even under these scenarios, and with confounding included, simulated early-life estimates remained smaller than the empirical effect estimates. When selection was instead specified to operate through mid-to-late adulthood adiposity, bias was expressed primarily in mid-to-late adulthood MR estimates, while early-life estimates remained close to the null. This pattern, consistent with the parental longevity analyses indicating survival-related differences are driven predominantly by mid-to-late adulthood adiposity, suggests that adulthood-driven selection does not account for the protective early-life MR effect estimates on breast cancer. Across extended simulations allowing more complex interaction structures, the joint empirical pattern of univariable and multivariable MR estimates was not reproduced under selection alone. Although the simulations were not parameterised to reproduce the exact selection structure of UK Biobank, parameters were intentionally varied beyond empirically typical magnitudes to evaluate whether extreme selection configurations could generate effect estimates comparable to those observed.

Taken together, these findings indicate that the selection mechanisms evaluated, while capable of inducing bias, do not adequately account for the magnitude and structure of the observed protective early-life MR effect estimate for breast cancer. More broadly, this work illustrates how triangulating complementary empirical analyses with targeted simulations can provide a practical approach for evaluating proposed selection mechanisms in lifecourse Mendelian randomization studies, rather than relying on any single diagnostic or sensitivity analysis. Alternative explanations for this inverse effect estimate have been proposed. In particular, higher adiposity during early life has been linked to differences in circulating sex hormone levels and lower mammographic breast density in adulthood, each of which has been implicated in breast cancer risk [5, 6]. Ongoing work aims to further clarify these pathways and the extent to which early-life adiposity may influence breast tissue development and hormonal exposures across the lifecourse.

## Data Availability

All code used to generate the simulations and reproduce the results reported here is publicly available at https://github.com/gracemarionpower/selection-bias-simulations/. The data underlying the findings can be derived directly from these simulations.

## Research Ethics and Informed Consent

The UK Biobank study has obtained ethics approval from the Research Ethics Committee (REC; approval number: 11/NW/0382). The UK Biobank study have obtained informed consent from all participants enrolled in UK Biobank. Empirical estimates were derived using data from the UK Biobank (app #81499). The remaining aspects of this study were based entirely on simulated data and did not involve human participants directly.

## CRediT authorship contribution statement

GMP: Conceptualization, Methodology, Software, Formal analysis, Investigation, Data curation; Writing - Original Draft, Writing - Review & Editing, Visualization; S: Methodology, Writing - Review & Editing; A : Methodology, Writing-Review & Editing; T R: Conceptualization, Formal analysis, Investigation, Data curation, Writing - Review & Editing; KT: Methodology, Writing - Review & Editing, Supervision; S: Conceptualization, Writing-Review & Editing, Supervision;GH: Conceptualization, Methodology, Software, Formal analysis, Investigation, Writing-Original Draft, Writing-Review & Editing, Visualization, Supervision.

## Data and computing code availability

All code used to generate the simulations and reproduce the results is publicly available at https://github.com/gracemarionpower/selection-bias-simulations/. Individual-level data used to derive these results can be obtained with an approved application to the UK Biobank study (http://www.ukbiobank.ac.uk). Data underlying the simulated analyses can be generated using the provided code.

