## Supplementary Material for "Triangulating evidence to evaluate selection bias in lifecourse Mendelian randomization studies: a practical framework illustrated by early-life adiposity and breast cancer"

### Supplementary materials

Table S8. Simulation results across 27 parameter scenarios of selection without interaction and interaction-based selection, under varying strengths of confounding... 19

#### Note S1: Data sources and variable definitions for empirical analyses

UK Biobank data were collected between 2006 and 2010 from individuals aged 40–69 years at baseline, using a prospective cohort design, and included clinical examinations, assays of biological samples, detailed self-reported health characteristics, and genome-wide genotyping [1]. Early-life adiposity was assessed via recall: adult participants reported whether, at age 10, they were "thinner," "about average," or "plumper" compared with peers. Adult adiposity was derived from clinically measured BMI (mean age 56.5 years) and recoded into a three-category variable ("thinner," "about average," "plumper") using the same proportions as the early-life measure [2, 3]. This was to ensure that derived effect estimates from subsequent analyses were as comparable as possible. Individuals that did not have data for both early-life and adult adiposity were excluded. Genetic variants strongly associated with early-life and adult adiposity (using $P < 5x10^{-8}$ and $r2 < 0.001$) were identified in a large-scale genome-wide association study (GWAS), previously undertaken on 463,005 individuals in the UK Biobank study, adjusting for age, sex, and genotyping chip[1, 4]. These instruments have been independently validated in three distinct cohorts, supporting their reliability in measuring early-life and adult adiposity[2, 3, 5]. Phenotypic data for breast cancer, were obtained from the UK Biobank. This was classified using the International Classification of Diseases, Tenth Revision (ICD-10) codes, described in Table S2. The UK Biobank study has obtained ethics approval from the Research Ethics Committee (REC; approval number: 11/NW/0382) and informed consent from all participants enrolled in UK Biobank. Estimates were derived using data from the UK Biobank (app #81499). Outcome GWAS summary statistics for parental survival were obtained from a published study of parental lifespan [6].

#### Table S1. Breast cancer outcome definition (ICD-10 codes)

| Measure | Definition | ICD-10 code(s) |
|---|---|---|
| Breast cancer | Malignant neoplasm of the breast | C500-06, C508-09 |

#### Note S2: Explaining proxy-MR results without the need for selection bias.

We adapted the simulation strategy described in Brumpton et al 2020 [7]. Briefly, we simulated nuclear families of two parents and two offspring, whereby parents each had independently sampled genotype values (i.e. no assortative mating) for 50 independent bi-allelic SNPs with allele frequencies sampled from a uniform distribution. Offspring inherited one allele randomly selected from each parent, independently from each SNP (i.e. no linkage between variants). Phenotypes were generated independently for each family member such that the 50 variants explained 40% of the variance in the exposure, exposure variance was fixed to 1, and the exposure influenced outcome liability with $l_i = -0.42x$ to match the empirical MR estimate, and a binary case control variable was generated sampling from a logistic transformation of the liability. Siblings were randomly assigned sex, and all males were set to be controls (i.e. no breast cancer). MR analysis was then performed by identifying genetic variants that were significant at p < 5e-8 in sibling 1, obtaining SNP-exposure effect estimates from sibling 1, and then performing logistic regression to obtain SNP-outcome effect estimates in female sibling 1 individuals (self), mothers, or female sibling 2 individuals (sibling). IVW analysis was performed on these summary statistics. For each simulation, 10000 families were simulated and the procedure was performed 100 times, each giving rise to three MR estimates (one for self, mother and sibling each).

### Figure S1: Explaining proxy-MR results without the need for selection bias.

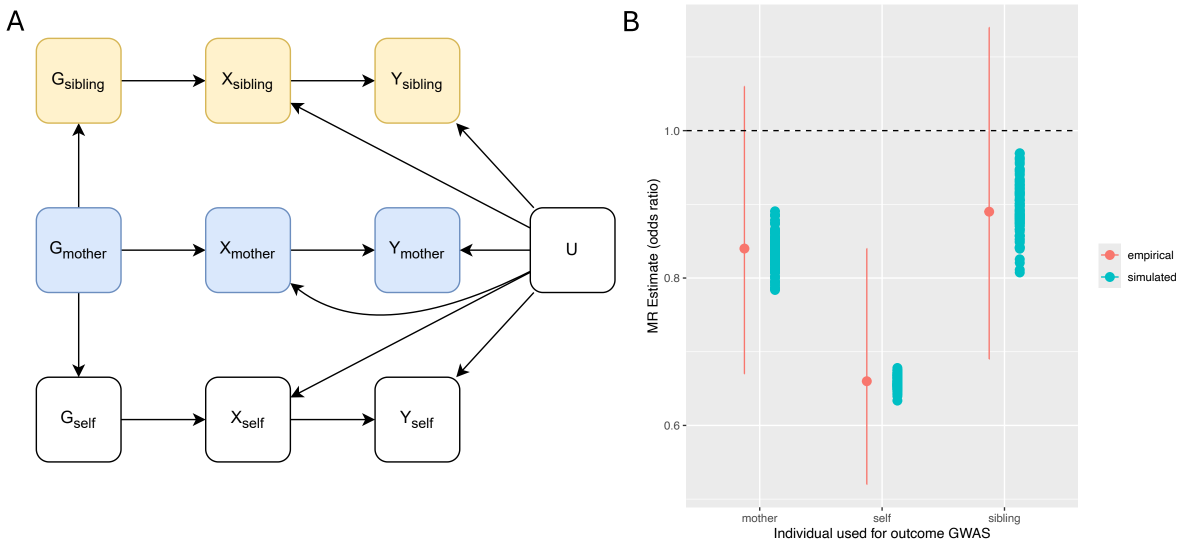

Schooling et al. observed that MR analysis of self-adiposity on sibling or mother breast cancer was attenuated compared to MR analysis of self-adiposity on self-breast cancer [8]. They proposed that this could be due to selection bias. Here we demonstrate that such a result is expected in the absence of selection bias. (A) the data generating mechanism that gives rise to (imperfectly) correlated outcomes amongst family members if risk factors have genetic instruments; (B) The empirical analysis of self-adiposity on self-breast cancer, mother breast cancer or sibling breast cancer (red points with 95% confidence intervals reported by Schooling et al) compared against corresponding simulation results drawn from the diagram in (A).

#### Note S2: MR analyses of parental survival to evaluate survival-related selection.

To further evaluate the plausibility of survival-related selection bias, we conducted an analysis using the early-life and mid-to-late adulthood adiposity instruments applied by Richardson et al. [5]. Parental survival was proxied using reported parental age at death (or current age if alive). Parental age at death strongly predicts participants' own lifespan at the population level [9, 10], yet it cannot be causally influenced by participants' early-life or mid-to-late adulthood adiposity. If the apparent protective effect estimate of increased early-life adiposity on breast cancer arose primarily from survival bias acting through the instruments, similar patterns would be expected for parental survival. Instead, higher mid-to-late adulthood adiposity showed a strong inverse effect on parental survival in both univariable (OR = 0.66, SE = 0.03, $P < 1\times10^{-37}$) and multivariable analyses (OR = 0.67, SE = 0.04, $P < 1\times10^{-21}$). Early-life adiposity also showed an inverse effect on parental survival in univariable analysis (OR = 0.80, SE = 0.03, $P < 1\times10^{-11}$), but this effect attenuated after adjusting for mid-to-late adulthood adiposity (OR = 1.00, SE = 0.05, P = 0.96) (Table S2). These findings suggest that survival differences are driven by mid-to-late adulthood, rather than early-life, adiposity and provide empirical reassurance that the inverse effect estimate of early-life adiposity on breast cancer is unlikely to be fully explained by survival bias. Effect estimates are presented as exponentiated MR coefficients (reported as ORs), derived from a survival-based outcome GWAS.

### Table S2. Mendelian randomization estimates of early-life and mid-to-late adulthood adiposity on parental longevity in UK Biobank.

Univariable and multivariable MR models were performed using genetic instruments for early-life and mid-to-late adulthood adiposity. Estimates are presented as odds ratios (OR) with standard errors (SE) and p-values (P).

| | | Univariable MR | | | Multivariable MR | | |
|---|---|---|---|---|---|---|---|
| Exposure | Outcome | OR | SE | P | OR | SE | P |
| Early-life adiposity | Parental longevity | 0.803 | 0.032 | 4.49E-12 | 0.997 | 0.048 | 9.55E-01 |
| Mid-to-late adulthood adiposity | Parental longevity | 0.663 | 0.032 | 1.66E-37 | 0.671 | 0.042 | 2.33E-21 |

#### Table S3. Empirical lifecourse Mendelian randomization results for the effect of early-life and mid-to-late adulthood adiposity on breast cancer risk

From Richardson et al., 2020 [5]

| Timepoint | Method | OR (95% CI) |
|---|---|---|
| Mid-to-late adulthood | Univariable | 0.82 (0.73, 0.92) |
| Early-life | Univariable | 0.63 (0.55, 0.72) |
| Mid-to-late adulthood | Multivariable | 1.08 (0.93, 1.27) |
| Early-life | Multivariable | 0.59 (0.50, 0.71) |

#### Note S3. Observational analysis as an empirical diagnostic for residual confounding.

We conducted a minimally adjusted logistic regression analysis using the UK Biobank observational dataset (female participants, n = 246,474) to assess the extent of confounding that might be present in the observed protective association between early-life adiposity and breast cancer risk as reported in empirical MR analyses. Early-life adiposity was modelled as the exposure and breast cancer status as the binary outcome, adjusting for age and mid-to-late adulthood adiposity. Adjustment for mid-to-late adulthood adiposity allowed estimation of the direct effect of early-life adiposity on breast cancer risk, analogous to the multivariable MR model, while age adjustment accounted for age-related variation in adiposity and breast cancer risk, acknowledging that additional covariate adjustment would not resolve the underlying confounding that motivates the use of MR. This analysis provided an empirical benchmark for the potential influence of confounding in observational data and informed whether inclusion of a confounder in the simulation models was warranted. Additional observational analyses were not central to our interpretation, as MR is employed precisely to mitigate confounding that cannot be resolved in conventional models.

Our real-data analysis acted as an empirical check on the direction and potential magnitude of confounding in conventional observational models. We fitted an observational logistic regression model among female UK Biobank participants only (n = 246,474), including early-life adiposity, mid-to-late adulthood adiposity, and age as covariates. Higher early-life adiposity was associated with a lower risk of breast cancer after adjustment for mid-to-late adulthood adiposity and age (logOR =-0.13, 95% CI:-0.15 to -0.11; OR = 0.88, 95% CI: 0.86 to 0.89) (Table S4). Notably, the magnitude of this association was substantially weaker than the corresponding MR estimates, indicating that confounding in observational data likely attenuates, rather than exaggerates, the protective effect. This empirical comparison motivated the inclusion of an unmeasured confounder in subsequent simulation analyses to evaluate how confounding and selection might jointly influence effect estimates.

Table S4. Observational association of early-life adiposity with breast cancer adjusted for age and mid-to-late adulthood adiposity

| Exposure | Outcome | logOR | 95% CI (logOR) | OR | 95% CI (OR) |
|---|---|---|---|---|---|
| Early-life adiposity | Breast cancer | -0.13 | -0.15,-0.11 | 0.88 | 0.86, 0.89 |

#### Note S5. Simulation framework for evaluating selection-induced collider bias.

For the simulations, we modelled early-life and mid-to-late adulthood adiposity as latent continuous traits. Latent continuous traits for early-life and mid-to-late adulthood adiposity were generated from genetic risk scores (GRS). Each GRS was simulated as a standard normal variable (mean zero, standard deviation one), with a genetic correlation of rg=0.67 between early-life and mid-to-late adulthood scores. Each GRS was specified to explain 10% of the variance in its respective adiposity trait. To capture the empirical observation that adiposity often persists across the lifecourse, mid-to-late adulthood adiposity was modelled to depend partly on early-life adiposity. Specifically, the latent adult adiposity included a direct contribution from the latent early-life adiposity (tracking parameter $\phi$=0.35), in addition to its own GRS and independent residual noise. This value was chosen with reference to estimates from three British birth cohorts, where median-quantile tracking coefficients between BMI at 11 and 42 years were reported in the range 0.28-0.53 across sexes and socioeconomic groups[11], consistent with a meta-analysis reporting an average correlation of ($r$ = 0.4) for early-life-to-mid-to-late adulthood BMI tracking[12]. In our model, however, $\phi$ represents only the direct carry-over from early-life to adulthofod, while additional correlation arises through genetic pathways; accordingly, we set $\phi$ at the lower end of the empirical range. This “tracking path” represents the propensity for children who are heavier than their peers to remain heavier in mid-to-late adulthood. All residuals were generated independently, and no further non-genetic correlation was introduced between early-life and mid-to-late adulthood adiposity.

The latent traits were then discretised into ordinal categories (0 = thinner, 1 = average, 2 = plumper) using thresholds corresponding to the 33rd and 84th percentiles of the UK Biobank distribution. Breast cancer status was assigned randomly using a fixed population prevalence of 1 in 7. We incorporated an unmeasured confounder to examine how additional dependence between the exposure and outcome might amplify collider bias.

Simulations used a sample size of n=246,511, chosen to reflect the number of female UK Biobank participants with available relevant data, and were repeated over 500 iterations. Within the selected sample, we estimated the direct effect of early-life adiposity on breast cancer risk, accounting for mid-to-late adulthood adiposity, using a two-stage residual inclusion (2SRI) logistic regression model. This two-exposure framework mirrors lifecourse MR designs, which jointly instrument early- and later-life adiposity to estimate the direct effect of early-life adiposity on disease risk, independent of adult adiposity[5]. From each model, we extracted the coefficient for early-life and adult adiposity to quantify the magnitude and direction of bias introduced under each selection scenario. In these simulations, breast cancer status was generated independently of adiposity, so the true causal effect of early-life and adult adiposity on breast cancer risk was set to zero; any estimated effect therefore reflects bias introduced by selection rather than genuine causality.

Selection into the analytic sample was modelled using a logistic function with no intercept (Eq. 1).

**Eq. 1**

$$\text{logit}(P(\text{selected})) = \beta_1 X_1 + \beta_2 X_2 + \beta_3 Y + \beta_4 (X_1 \times Y)$$

where:

- $X_1$: early-life adiposity
- $X_2$: mid-to-late adulthood adiposity
- $Y$: binary breast cancer status
- $\beta_1$: additive effect of early-life adiposity on the probability of being selected
- $\beta_2$: additive effect of adult adiposity (fixed at zero)
- $\beta_3$: additive effect of breast cancer status on selection
- $\beta_4$: interaction effect between early-life adiposity and breast cancer status, representing differential selection probabilities according to joint values of exposure and outcome

Because the intercept was omitted, the baseline selection probability is arbitrary; coefficients are therefore interpreted as relative selection effects rather than calibrated to reproduce the absolute UK Biobank participation rate.

The model of Equation (1) defined the data-generating process for selection probability in all simulations. Parameters $\beta_1$, $\beta_3$, and $\beta_4$ directly determined the likelihood of inclusion, rather than assuming a constant participation rate. Negative $\beta_1$ values represented preferential inclusion of thinner children; negative $\beta_3$ values modelled under-representation of cancer cases; and negative $\beta_4$ values represented compounded exclusion of individuals who were both heavier in early-life and developed breast cancer (Table S5). Parameter values were specified to span no selection and progressively increasing magnitudes of selection effects. The highest-magnitude values were included to evaluate whether extreme selection configurations could generate bias comparable to the observed MR estimates under a null causal model.

#### Table S5. Interpretation of selection parameters used in simulations.

| Mechanism | Parameter | Level | Selection-odds multiplier | Interpretation |
|---|---|---|---|---|
| Body-size selection[&] | $\beta_1$ | 0.00 | 1.00 | No body-size selection; participants of all early-life adipositys included equally. |
| | | -0.25 | 0.78 per step | Intermediate-magnitude selection favouring thinner participants; each one-category increase reduces inclusion odds by 22%. |
| | | -0.50 | 0.61 per step | High-magnitude selection favouring thinner participants; each one-category increase reduces inclusion odds by 39%. |
| Breast-cancer under-selection | $\beta_3$ | 0.00 | 1.00 (case vs non-case) | No case under-selection; breast-cancer cases and non-cases included equally. |
| | | -0.25 | 0.78 (case vs non-case) | Intermediate-magnitude under-selection of breast-cancer cases; cases have 22% lower odds of inclusion than non-cases. |
| | | -0.50 | 0.61 (case vs non-case) | High-magnitude under-selection of breast-cancer cases; cases have 39% lower odds of inclusion than non-cases. |
| Interaction-dependent selection† | $\beta_4$ | 0.00 | 1.00 | No interaction; adiposity and case status influence selection independently. |
| | | -0.25 | 0.78 per step (cases only) | Intermediate-magnitude interaction; heavier breast-cancer cases are additionally less likely to be included, beyond $\beta_1$ and $\beta_3$. |
| | | -0.50 | 0.61 per step (cases only) | High-magnitude interaction; compounded exclusion of breast-cancer cases who were heavier in early-life, beyond $\beta_1$ and $\beta_3$. |

[&] We modelled early-life adiposity as exerting a linear effect on the logit of the probability of selection, with optional modification by breast cancer status. This specification assumes that each step heavier in adiposity is associated with a constant proportional change in the odds of being included. In reality, the relation between adiposity and study participation may be non-linear. For example, both very thin and very heavy children may be disproportionately excluded and these patterns may interact with disease status. Such complexities were not incorporated here, as the objective was to evaluate simple, interpretable selection mechanisms and their capacity to generate upper-bound collider bias; they represent important extensions for future work.† The $\beta_4$ multiplier applies in addition to $\beta_1$ and $\beta_3$ and only to breast-cancer cases; e.g., at $\beta_1 = \beta_3 = \beta_4 = -0.50$, a one-category heavier case has inclusion odds $0.61 \times 0.61 \times 0.61 \approx 0.23$ of the reference.

This specification captures two theoretically distinct but potentially co-occurring mechanisms of collider bias (Table S6). Selection without interaction refers to situations where early-life adiposity and breast cancer status independently affect inclusion in the analytical sample. In contrast, interaction-based selection reflects differential inclusion odds according to joint values of exposure and outcome, such that exclusion is amplified for individuals who were both heavier in early-life and developed breast cancer.

We evaluated all 27 combinations of $\beta_1$, $\beta_3$, and $\beta_4$ across none, moderate, and strong levels of selection, spanning settings with no selection bias, selection without interaction bias, and interaction-driven bias.

#### Table S6. Conceptual mechanisms of selection bias modelled in simulations.

| Mechanism | Description | Example contexts |
|---|---|---|
| Selection without interaction | Early-life adiposity and breast cancer status independently affect inclusion in the analytical sample. | Heavier children lost to follow-up due to long-term health conditions or socioeconomic disadvantage; breast cancer cases underrepresented due to earlier mortality or lower participation. |
| Interaction-based selection | Inclusion depends jointly on early-life adiposity and breast cancer status, such that exclusion is amplified for heavier individuals who also develop breast cancer. | Heavier children who later develop breast cancer less likely to be recruited due to compounded health risks, earlier death, or structural disadvantage. |

#### Note S6. Magnitude of interaction coefficients influencing selection.

Using incidence rates from Fry et al (2017) [13] for various cancers in UK Biobank participants ($\mathrm{I_{ukb}}$) and the general UK population ($\mathrm{I_{pop}}$), we made approximate estimates of the relationship between disease status and sample selection ($\mathrm{RR} = \mathrm{I_{ukb}}/\mathrm{I_{pop}}$). As the Fry et al. (2017) study also stratified these incidence rates by age, we were able to calculate approximate estimates of the influence of the disease status x age interaction based on $\mathrm{RR_{sel,int}} = \mathrm{RR_{sel,age=45-49}}/\mathrm{RR_{sel,age=70-74}}$.

Figure S2. Risk ratio for selection into UK Biobank for different cancers and stratified by age and sex.

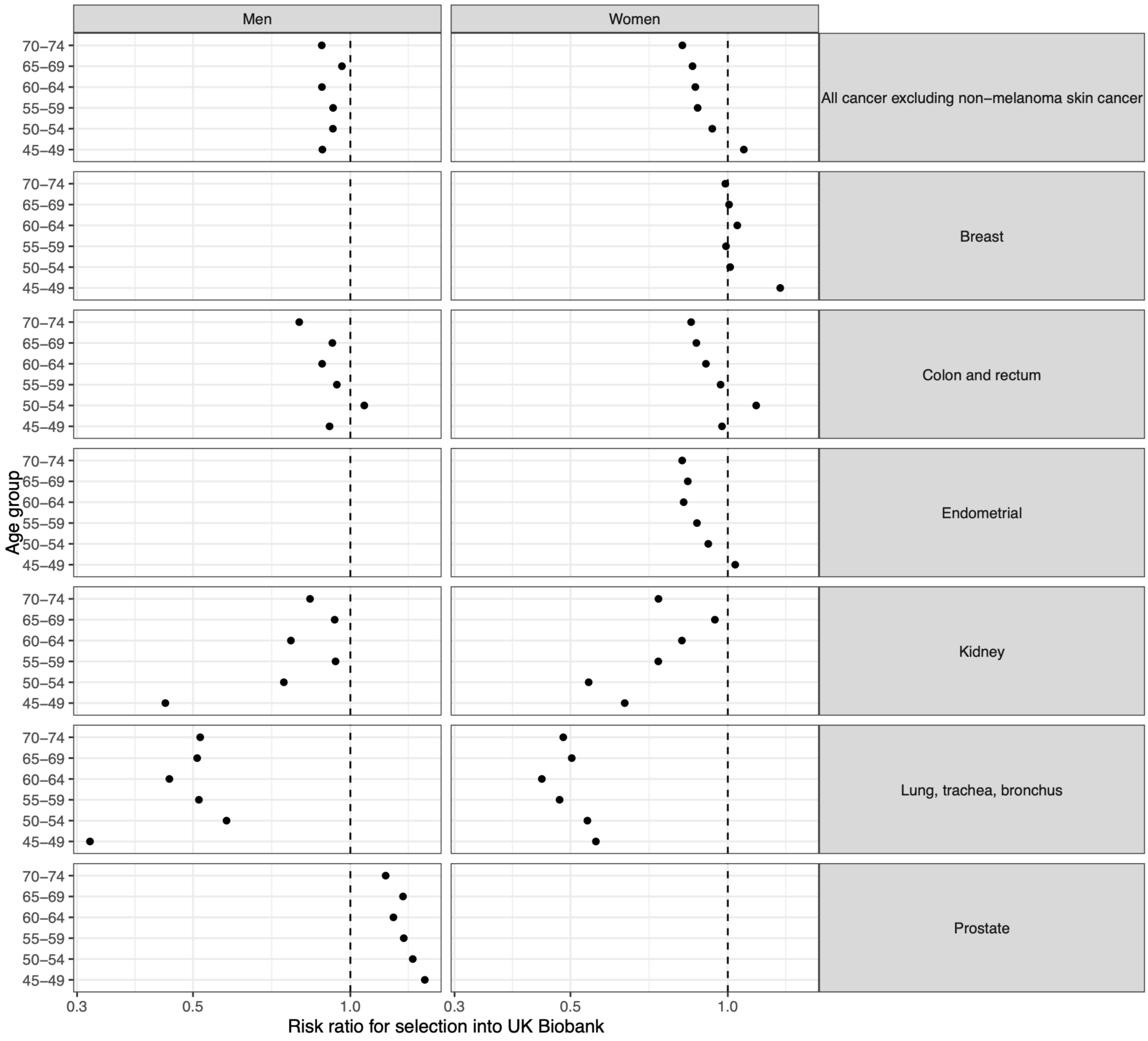

Table S7: Disease x age interactions on sample selection.

| Cancer type | sex | $RR_{sel}$ | $RR_{sel,int}$ |
|---|---|---|---|
| All cancer excluding non-melanoma skin cancer | men | 0.88 | 1.00 |
| All cancer excluding non-melanoma skin cancer | women | 0.94 | 0.76 |
| Breast | women | 1.12 | 0.79 |
| Colon and rectum | men | 0.85 | 0.88 |
| Colon and rectum | women | 0.91 | 0.87 |
| Endometrial | women | 0.92 | 0.79 |
| Kidney | men | 0.61 | 1.89 |
| Kidney | women | 0.68 | 1.16 |
| Lung, trachea, bronchus | men | 0.40 | 1.63 |
| Lung, trachea, bronchus | women | 0.52 | 0.87 |
| Prostate | men | 1.27 | 0.84 |

### Table S8. Simulation results across 27 parameter scenarios of selection without interaction and interaction-based selection, under varying strengths of confounding.

For each scenario, we report the mean estimated log odds ratio (logOR) for early-life adiposity, its standard deviation (SD), and the mean standard error (SE) across 500 replicates. Equivalent columns are also reported for adult adiposity estimates. The confounding status column indicates whether a latent confounder (U) influencing both adiposity and breast cancer risk was included in the data-generating model (mild or strong).

| β1 - child adiposity selection | β3 - breast cancer selection | β4 - interaction selection | Mean logOR (Child) | SD logOR (Child) | Mean SE (Child) | Mean logOR (Adult) | SD logOR (Adult) | Mean SE (Adult) | Confounding status |
|---|---|---|---|---|---|---|---|---|---|
| -0.50 | -0.50 | -0.50 | -0.367 | 0.092 | 0.087 | -0.001 | 0.072 | 0.068 | Mild confounding |
| -0.50 | -0.50 | -0.25 | -0.222 | 0.088 | 0.087 | -0.002 | 0.071 | 0.068 | Mild confounding |
| -0.50 | -0.50 | 0.00 | -0.079 | 0.078 | 0.087 | 0.003 | 0.063 | 0.068 | Mild confounding |
| -0.50 | -0.25 | -0.50 | -0.314 | 0.083 | 0.082 | -0.003 | 0.066 | 0.064 | Mild confounding |
| -0.50 | -0.25 | -0.25 | -0.185 | 0.078 | 0.082 | 0.001 | 0.061 | 0.064 | Mild confounding |
| -0.50 | -0.25 | 0.00 | -0.046 | 0.078 | 0.081 | 0.002 | 0.060 | 0.063 | Mild confounding |
| -0.50 | 0.00 | -0.50 | -0.255 | 0.085 | 0.078 | -0.004 | 0.064 | 0.060 | Mild confounding |
| -0.50 | 0.00 | -0.25 | -0.132 | 0.078 | 0.077 | -0.009 | 0.063 | 0.060 | Mild confounding |
| -0.50 | 0.00 | 0.00 | -0.019 | 0.076 | 0.077 | 0.004 | 0.059 | 0.060 | Mild confounding |
| -0.25 | -0.50 | -0.50 | -0.347 | 0.076 | 0.085 | 0.008 | 0.063 | 0.067 | Mild confounding |
| -0.25 | -0.50 | -0.25 | -0.19 | 0.082 | 0.084 | -0.004 | 0.065 | 0.067 | Mild confounding |
| -0.25 | -0.50 | 0.00 | -0.03 | 0.084 | 0.084 | -0.007 | 0.068 | 0.067 | Mild confounding |
| -0.25 | -0.25 | -0.50 | -0.295 | 0.078 | 0.079 | -0.006 | 0.063 | 0.063 | Mild confounding |
| -0.25 | -0.25 | -0.25 | -0.172 | 0.072 | 0.079 | 0.006 | 0.059 | 0.063 | Mild confounding |
| -0.25 | -0.25 | 0.00 | -0.019 | 0.075 | 0.079 | -0.003 | 0.065 | 0.063 | Mild confounding |
| -0.25 | 0.00 | -0.50 | -0.249 | 0.075 | 0.075 | -0.005 | 0.059 | 0.060 | Mild confounding |
| -0.25 | 0.00 | -0.25 | -0.132 | 0.077 | 0.075 | -0.002 | 0.059 | 0.060 | Mild confounding |
| -0.25 | 0.00 | 0.00 | -0.006 | 0.071 | 0.074 | 0.000 | 0.057 | 0.060 | Mild confounding |
| 0.00 | -0.50 | -0.50 | -0.311 | 0.083 | 0.083 | -0.004 | 0.069 | 0.067 | Mild confounding |
| 0.00 | -0.50 | -0.25 | -0.157 | 0.080 | 0.082 | -0.001 | 0.068 | 0.067 | Mild confounding |
| 0.00 | -0.50 | 0.00 | -0.003 | 0.083 | 0.082 | 0.000 | 0.063 | 0.067 | Mild confounding |
| 0.00 | -0.25 | -0.50 | -0.288 | 0.081 | 0.077 | 0.002 | 0.064 | 0.063 | Mild confounding |
| 0.00 | -0.25 | -0.25 | -0.141 | 0.079 | 0.077 | 0.001 | 0.063 | 0.063 | Mild confounding |
| 0.00 | -0.25 | 0.00 | 0.001 | 0.079 | 0.076 | -0.005 | 0.064 | 0.062 | Mild confounding |
| 0.00 | 0.00 | -0.50 | -0.251 | 0.071 | 0.073 | -0.005 | 0.061 | 0.059 | Mild confounding |
| 0.00 | 0.00 | -0.25 | -0.125 | 0.074 | 0.073 | -0.002 | 0.062 | 0.059 | Mild confounding |
| 0.00 | 0.00 | 0.00 | -0.005 | 0.070 | 0.072 | -0.002 | 0.060 | 0.059 | Mild confounding |
| -0.50 | -0.50 | -0.50 | -0.421 | 0.092 | 0.096 | -0.012 | 0.071 | 0.074 | Strong confounding |
| -0.50 | -0.50 | -0.25 | -0.258 | 0.101 | 0.095 | -0.017 | 0.078 | 0.073 | Strong confounding |
| -0.50 | -0.50 | 0.00 | -0.102 | 0.095 | 0.094 | -0.018 | 0.069 | 0.073 | Strong confounding |
| -0.50 | -0.25 | -0.50 | -0.372 | 0.090 | 0.090 | -0.009 | 0.067 | 0.069 | Strong confounding |
| -0.50 | -0.25 | -0.25 | -0.226 | 0.093 | 0.089 | -0.008 | 0.072 | 0.069 | Strong confounding |

| | | | | | | | | | |
|---|---|---|---|---|---|---|---|---|---|
| -0.50 | -0.25 | 0.00 | -0.079 | 0.088 | 0.088 | -0.009 | 0.071 | 0.069 | Strong confounding |
| -0.50 | 0.00 | -0.50 | -0.317 | 0.088 | 0.085 | -0.011 | 0.069 | 0.066 | Strong confounding |
| -0.50 | 0.00 | -0.25 | -0.177 | 0.089 | 0.085 | -0.016 | 0.069 | 0.065 | Strong confounding |
| -0.50 | 0.00 | 0.00 | -0.045 | 0.091 | 0.084 | -0.016 | 0.069 | 0.065 | Strong confounding |
| -0.25 | -0.50 | -0.50 | -0.382 | 0.087 | 0.092 | -0.006 | 0.074 | 0.073 | Strong confounding |
| -0.25 | -0.50 | -0.25 | -0.225 | 0.096 | 0.091 | -0.010 | 0.075 | 0.072 | Strong confounding |
| -0.25 | -0.50 | 0.00 | -0.056 | 0.090 | 0.091 | -0.013 | 0.069 | 0.072 | Strong confounding |
| -0.25 | -0.25 | -0.50 | -0.332 | 0.088 | 0.086 | -0.010 | 0.070 | 0.068 | Strong confounding |
| -0.25 | -0.25 | -0.25 | -0.19 | 0.083 | 0.086 | -0.016 | 0.066 | 0.068 | Strong confounding |
| -0.25 | -0.25 | 0.00 | -0.043 | 0.088 | 0.085 | -0.006 | 0.068 | 0.067 | Strong confounding |
| -0.25 | 0.00 | -0.50 | -0.296 | 0.089 | 0.082 | -0.008 | 0.071 | 0.065 | Strong confounding |
| -0.25 | 0.00 | -0.25 | -0.155 | 0.085 | 0.081 | -0.011 | 0.065 | 0.064 | Strong confounding |
| -0.25 | 0.00 | 0.00 | -0.027 | 0.081 | 0.080 | -0.010 | 0.063 | 0.064 | Strong confounding |
| 0.00 | -0.50 | -0.50 | -0.33 | 0.095 | 0.089 | -0.012 | 0.076 | 0.072 | Strong confounding |
| 0.00 | -0.50 | -0.25 | -0.169 | 0.086 | 0.088 | -0.011 | 0.069 | 0.071 | Strong confounding |
| 0.00 | -0.50 | 0.00 | -0.014 | 0.086 | 0.088 | -0.006 | 0.070 | 0.071 | Strong confounding |
| 0.00 | -0.25 | -0.50 | -0.3 | 0.088 | 0.084 | -0.013 | 0.071 | 0.068 | Strong confounding |
| 0.00 | -0.25 | -0.25 | -0.15 | 0.089 | 0.083 | -0.014 | 0.071 | 0.067 | Strong confounding |
| 0.00 | -0.25 | 0.00 | -0.015 | 0.086 | 0.082 | -0.010 | 0.072 | 0.066 | Strong confounding |
| 0.00 | 0.00 | -0.50 | -0.277 | 0.082 | 0.079 | -0.004 | 0.067 | 0.064 | Strong confounding |
| 0.00 | 0.00 | -0.25 | -0.138 | 0.079 | 0.078 | -0.009 | 0.061 | 0.063 | Strong confounding |
| 0.00 | 0.00 | 0.00 | -0.012 | 0.083 | 0.078 | -0.005 | 0.066 | 0.063 | Strong confounding |

#### Figure S3. Coverage of the empirical MR estimate across selection scenarios in the absence of confounding.

Heatmap displaying the proportion of simulation replicates in which the empirical MR estimate (logOR =-0.53) was contained within the 95% confidence interval. Columns represent levels of interaction-dependent selection, rows represent levels of breast cancer selection, and the x-axis shows early-life adiposity selection. Cell shading reflects coverage percentage, from 0% (white) to 100% (dark blue).

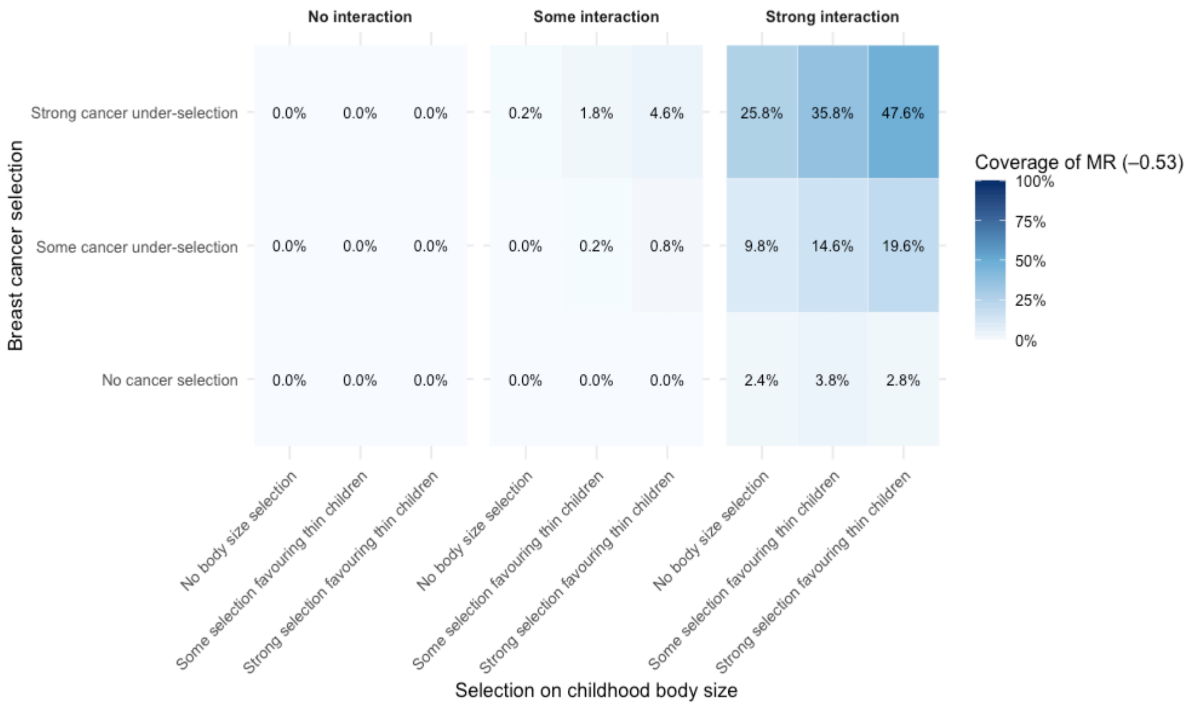

#### Figure S4. Simulated bias under selection acting on mid-to-late adulthood adiposity.

Mean log(OR) estimates for mid-to-late adulthood (dashed) and early-life (solid) adiposity on breast cancer across 500 simulation replicates are shown under increasing strengths of unmeasured confounding (U) affecting both adiposity and breast cancer risk. Horizontal panels represent different confounding strengths (none, mild, strong), and vertical panels represent increasing levels of interaction-driven selection acting on mid-to-late adulthood adiposity. Error bars indicate the 2.5th-97.5th percentile range across replicates. The red dashed line marks the observed mid-to-late adulthood MR estimate (logOR =-0.53

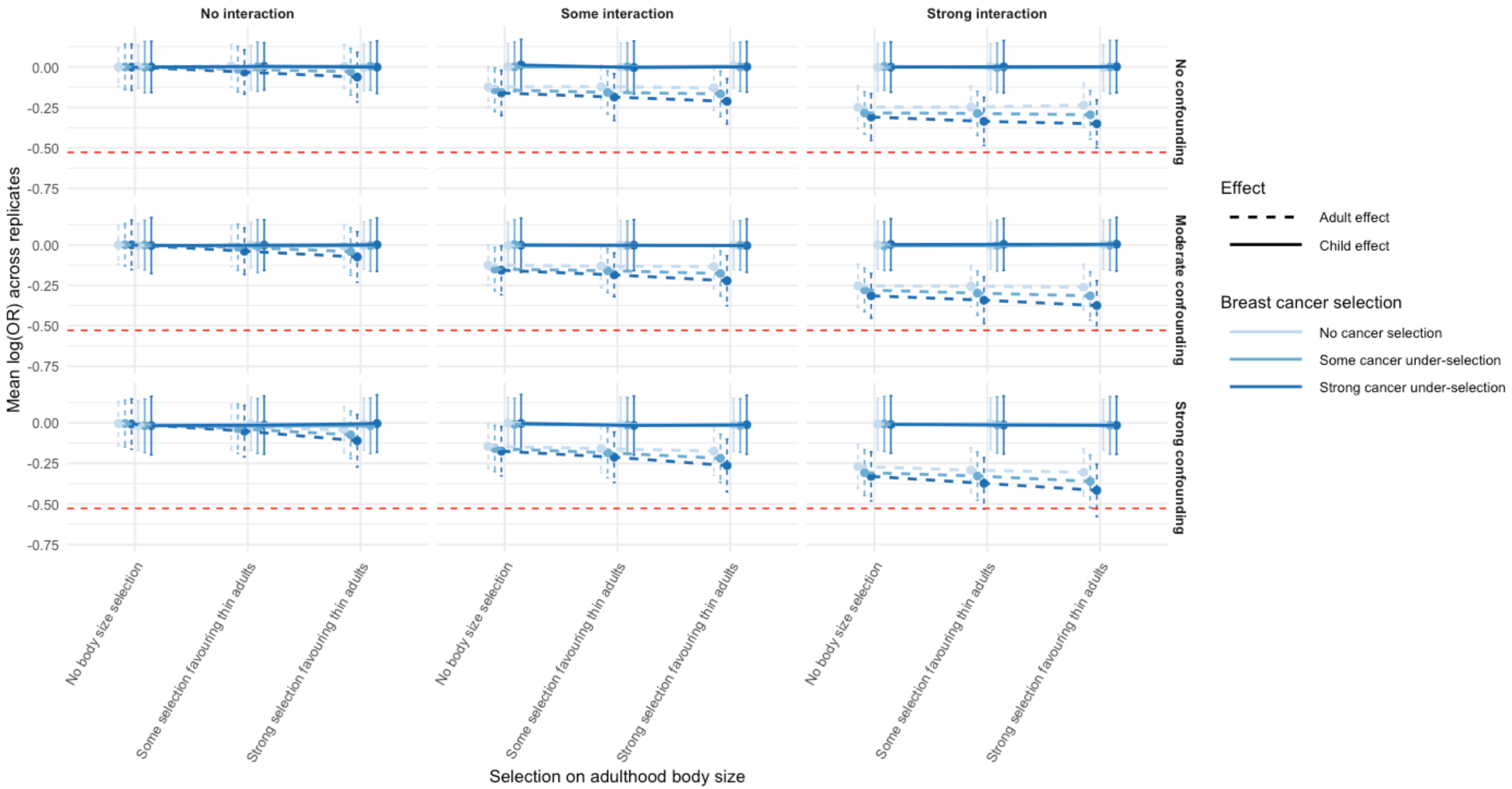

#### Note S7. Recapitulating univariable and multivariable MR patterns under simulated selection mechanisms

Next, we extended the selection model to explore how selection could explain the broader pattern of univariable and multivariable results observed empirically (Eq. 2). Our objective is to find examples of selection parameters that lead to all 4 empirical estimates in Table S3 (Richardson et al.[5]) being matched in the simulation. i.e. We are seeking data generating models that would lead to univariable estimates of early-life and mid-to-late adulthood adiposity being both strongly negatively associated, but in the multivariable analysis the mid-to-late adulthood effect estimate attenuates to the null. Our revised

Eq. 2

$$logit(P(selected)) = \beta_1 X_1 + \beta_2 X_2 + \beta_3 Y + \beta_4(X_1 \times Y) + \beta_5(X_2 \times Y) + \beta_6(X_1 \times X_2)$$
$$\beta_7(X_1 \times X_2 \times Y) + \beta_8 U$$

where:

- $\beta_5$: interaction effect between mid-to-late adulthood adiposity and breast cancer status, representing differential selection probabilities according to joint values of exposure and outcome
- $\beta_6$: interaction effect between early-life adiposity and mid-to-late adulthood adiposity, for example capturing if individuals who were larger in both early-life and mid-to-late adulthood were more likely to be excluded
- $\beta_7$: interaction effect between early-life adiposity, mid-to-late adulthood adiposity and breast cancer status, representing differential selection probabilities according to joint values of exposure and outcome
- $\beta_8$: The influence of the unmeasured confounder on selection. Note that the unmeasured confounder has influences on cancer outcome, early-life adiposity and mid-to-late adulthood adiposity

For these simulations we allowed all selection coefficients to vary across the range (0, -0.1, ..., -0.6). We estimated four effects: (i) the univariable effect of early-life adiposity on breast cancer, (ii) the univariable effect of mid-to-late adulthood adiposity on breast cancer, (iii) the multivariable direct effect of early-life adiposity on breast cancer (adjusting for mid-to-late adulthood adiposity), and (iv) the multivariable direct effect of mid-to-late adulthood adiposity on breast cancer (adjusting for early-life adiposity).

To examine whether the selection mechanism of a particular simulation could explain the empirical results, for each of the four estimates (j) we estimated the probability $p_j$ that they overlapped the corresponding empirical estimate using

$$z_j = abs(\hat{\beta}_{sim,j} - \hat{\beta}_{empirical,j})/\sqrt{\sigma^2_{sim,j} + \sigma^2_{empirical,j}}\,,\; p_j = P(|Z| > z_j), Z \sim N(0,1).$$

where $\hat{\beta}_{sim,j}$ is the simulated estimate for effect $\mathrm{j}$ with standard error $\sigma^2_{sim,j}$, with $\hat{\beta}_{empirical,j}$ and $\sigma^2_{empirical,j}$ the corresponding values from the empirical analysis in Table S3. For simplicity, we counted a simulation estimate as overlapping the empirical estimate if it overlap probability $\mathrm{p_j} > 0.1$. A 'successful' combination of parameters would give rise to all four values of $\mathrm{p_j} > 0.1$.
